# Modeling and Simulation: A study on predicting the outbreak of COVID-19 in Saudi Arabia

**DOI:** 10.1101/2021.01.17.21249837

**Authors:** Ahmed Msmali, Zico Mutum, Idir Mechai, Abdullah Ahmadini

## Abstract

The novel coronavirus (Covid-19) infection has resulted in an ongoing pandemic affecting health system and economy of more than 200 countries around the world. Mathematical models are used to predict the biological and epidemiological trends of an epidemic and develop methods for controlling it. In this work, we use mathematical model perspective to study the role of behavior change in slowing the spread of the COVID-19 disease in Saudi Arabia. The real-time updated data from 1^st^ May 2020 to 8^th^ January 2021 is collected from Saudi Ministry of Health, aiming to provide dynamic behaviors of the pandemic in Saudi Arabia. During this period, it has infected 297,205 people, resulting in 6124 deaths with the mortality rate 2.06 %. There is weak positive relationship between the spread of the infection and mortality (*R*^2^ =0.412). We use Susceptible-Exposed-Infection-Recovered (SEIR) mode, the logistic growth model and with special focus on the exposed, infection and recovery individuals to simulate the final phase of the outbreak. The results indicate that social distancing, good hygienic conditions, and travel limitation are the crucial measures to prevent further spreading of the epidemic.

## 1. Introduction

Coronavirus are the large family of virus that are spread among humans, animals and livestock. The virus may spread from birds such as bats to human through various intermediate host and cause severe respiratory syndrome [7]. They cause generally mild respiratory infection, dry cough, fever, and difficult of breathing. During the month of November, the first coronavirus outbreak has occurred in Wuhan, the capital city of the Hubei province and the seventh largest city of China [2-4]. In 31 December 2019, the first reported cases of the infected persons in Wuhan city were 25, including 7 critical ill cases. Since then, the epidemic outbreak has drawn a serious concern and global attention. The Chinese authorities on 7 January 2020, has identified the novel coronavirus as the causative agent. However, on 10 January 2020, the World Health Organization (WHO) has designated the novel coronavirus as 2019-nCoV. On the same day, the WHO released a wide range of interim guidance for all countries on how to get prepare for and responding to COVID-19 pandemic. This includes strategy and plan on how to monitor for potentially infected people, collect and test samples. A practical guidance on how to manage patients, control and mitigate the burden generated by the infection in health centers. This guidance also highlights the maintenance of right drug supplies and effectively communicate with the lay public regarding the new virus [5]. On 12 February 2020, the WHO has re-named the 2019-nCoV as sever acute respiratory syndrome coronavirus 2 (SARS-CoV-2). It has been announced that the official name of the virus responsible for causing coronavirus disease is SARS-CoV-2. The World Health Organization officially declared the outbreak as a global pandemic as the new coronavirus has rapidly spread to all over around 220 countries around the world [1].

On 2 March 2020, the first case of coronavirus in Saudi Arabia was identified and announced by Ministry of Health. From the first week of March, the number of confirmed coronavirus COVID-19 cases are gradually increasing and has led to 2932 confirmed cases in Saudi Arabia as of April 9, 2020. The pick time of increasing the infection cases were noticed in June and July months. At present the new cases of infection is decreasing gradually. On 8^th^ January 2021 the total number of 363061 confirmed cases of COVID-19 infection with 354,443 recovered and 6246 deaths have been conformed in Saudi Arabia (shown in Figure 1).

**FIGURE 1:**
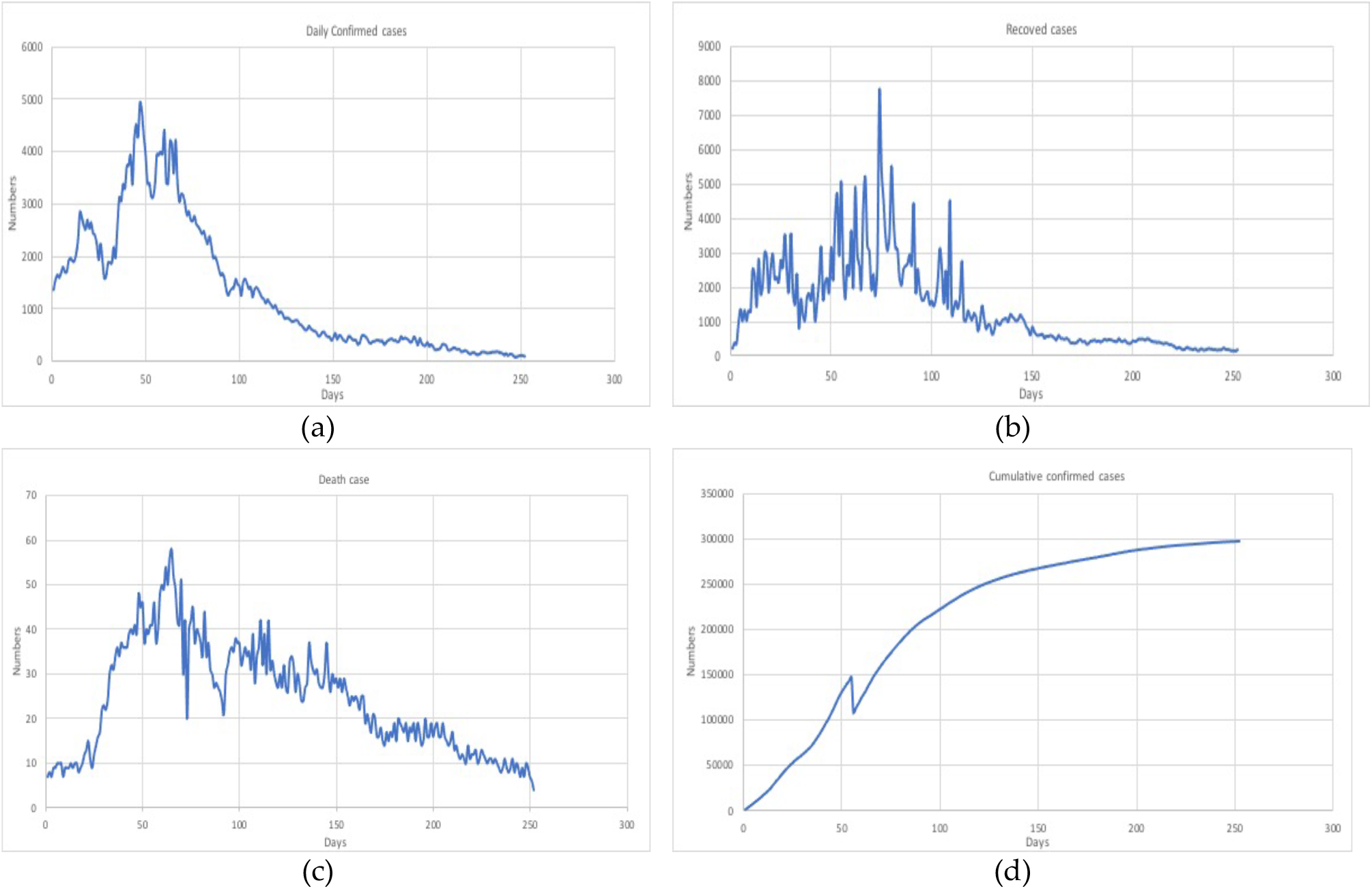
Number of (a) confirmed, (b) recovered (c) death cases and (d) cumulative confirmed cases due to COVID-19 in Saudi Arabia from 1^st^ May 2020 to 8^th^ January 2021.

Mathematical modeling and simulations are used as an important tool to predict the possibility and severity of disease outbreak and provide information to understand its dynamic behavior of the infection. Further, mathematical models have been widely used as an essential tool for investigating the dynamics of the spread of infectious diseases such as Foot-and-Mouth Disease, SARS, Ebola and Zika [10],[11],[12],[13].This article aims to further understand the transmission dynamics of the virus infection and facilitate the control measures using the real data from 1^st^ March 2020 to 8^th^ January 2021. We study the basic reproduction number of the virus at the current outbreak. It is important to predict the pandemic trend of COVID-19 in Saudi Arabia to help the strategies of control and prevention measures taken by the Ministry of health.

## 2. Related works

Adiga et al. [14] used both Auto Regressive Integrated Moving Average (ARIMA) and logistic growth models to study the trend and to provide short and long-term forecasting of the prevalence of COVID-19 cases and dynamics. The main idea of the ARIMA model is to precisely estimate the short-term prevalence of recovered and death cases due to infection in Saudi Arabia. The result of the model was outstanding with regards to mean absolute percentage error, mean absolute percentage error, mean absolute error and coefficient of determination. It has more advantage in short-term forecasting which gives more accurate projections of the pandemic. For the long-term forecasting they used logistic growth model.

Batista, M. [26] estimated the final size and its peak time of coronavirus COVID-19 in China, South Korea, and the rest of the world by using logistical growth regression model. According to the model prediction, the daily epidemic size forecasts begin to converge, and concluded that the outbreak is under control.

Ashleign et al. [15] proposed an aged-structured compartmental model of COVID-19 transmission in the population of Ontario, Canada. The model was stratified by age groups and used modified susceptible-exposed-infectious-recovered incorporating additional compartments for public health, severities of clinical symptoms and risk of hospital admission.

Chen et al., [16] developed a Bats-Hosts-Reservoir-People (BHRP) transmission network model to simulate the infection probability from bats to human. Further, they simplified the model as reservoir-people (RP) transmission. The estimated result of basic reproduction number was 2.30. And the simulation results showed that it was 3.58 from person to person. The results indicated that infection rate for coronavirus in some countries might be higher than MERS.

Din et al., [17] used SIR model showed that some factors such as exposure, death, and cure rates affect the susceptible, infected and recovered population involving immigration. The simulated results for infection rate increase with involving the concern immigration rate. And with smaller immigration rate the infection slows as compare at higher over and vice versa.

When the novel COVID-19 started to spread in China, many researchers discussed applying different models in integer, stochastic and in fractional order. Thabet et al., [19] modeled an existence results using Atangana–Baleanu–Caputo (ABC) derivative with fractional order. Further, simulation is performed using Adams Bashforth numerical technique concerning results for two famous cities of China known as Wuhan and Huanggang. Fractional models are more likely to be accurate in predicting the infection dynamics.

The SIR model with stochastic approaches are likely to produce the results different from deterministic models whenever the model is run for some parameters. Zhang et al., [18] study the effects of the environment on spreading of COVID-19 using stochastic modeling. The threshold of the stochastic COVID- 19 model is determined according to the large and small noise.

Meo et al., [20] investigated the biological and epidemiological trends in the prevalence and mortality due to outbreaks of COVID-19 infection. The data of the outbreak were obtained from World Health Organization. The study is done on gender-bases analysis demonstrates that the identified cases consisted mostly of men with a median age range of 50-65 years. And concluded that there was a large variation in the biological and epidemiological trends both in growth factor of number of cases and the mortality cases.

The nature of the spread of this pandemic in each country follows almost the same scenario. At the first phase, the number of infected people increases exponentially and then its growth slows down. A number of mathematical models of different complexity levels are used to predict the dynamics of the spread of COVID-19. The logistic model describes properly the growth rate of infected people by coronavirus and fitted with the available data. The use of a generalized logistic growth model improves the agreement with the real data, however, prognostic number of the total number of cases is slightly higher than the simple logistic model [33].

Wu et al., [23] employ for the classical logistic growth model and generalized form of the model to compare the outbreak dynamics in the 29 provinces in China. The model estimates growth rate of infection decays as fewer susceptible people are available for infection and better prediction at the later stage of the epidemic.

Alrasheed et al., [24] built a network simulation model of the spread of COVID-19 in Saudi Arabia using SIR model. A set of attributes such as age, gender, nationality and location are defined for nodes in network simulation. They also computed the percentage of people that must be vaccinated to stop the epidemic.

Adekunle et al., [21] analyses various forms of mathematical models that are relevant for the containment, risk analysis, and features of COIVD-19. The SEIR model seems the most reliable extension model because of its plausibility in explaining heterogeneous changes in behavior. And concluded, that mathematical models can significantly guide the implementation of public health decision when properly expressed and estimated.

## 3. Materials and Methods

The study of this research work is conducted in the department of mathematics, college of science, Jazan University, Jazan, Saudi Arabia. We collected the real-life data on based on the number of confirmed cases, recovered cases and number of deaths due to coronavirus COVID-19 infections all over the regions of Saudi Arabia. The cumulative data are collected and obtained from the World Health Organization (WHO) and Ministry of Health, Saudi Arabia (Covid19 Command and Control Center CCC,). We also obtained data from search engines including [1] and research articles published in [8]. We access the reproduction number which represents the average number of people to which an infected person can transmit the virus. Preliminary studies had estimated the reproduction number to be between 1.5 and 3.5. The spread of the virus will disappear gradually with a reproduction number below1. As for comparison, the reproduction number for common flu is 1.3 and SARS is 2.0. [1].

### 3.1 Ethical consideration

In our study, the information on epidemiological trends and current situation of coronavirus COVID 19 infection is taken from the World Health Organization, Worldmeter-Coronavirus and Ministry of Health, Covid19 Command and Control Center CCC. So, ethical approval is not required.

### 3.2 Statistical Data and Analysis

The updated data related to cumulated infected cases and recovery are analyzed. The Table 1, shows the number of confirmed and death cases at seven major cities in Saudi Arabia. The regression analysis is performed using the data from the Table 1. We found that the coefficient of determination *R*^2^ is 0.34 and a p-value is .00014, which is considered significant. According to the population in each city, the city of Dhahran has the highest percentage of confirmed (average 4.5%) and death (average 0.8%) cases. The city of Tabuk has the lowest percentage of confirmed (average 0.89%) and death (average 0.018%) cases.

**TABLE 1:**
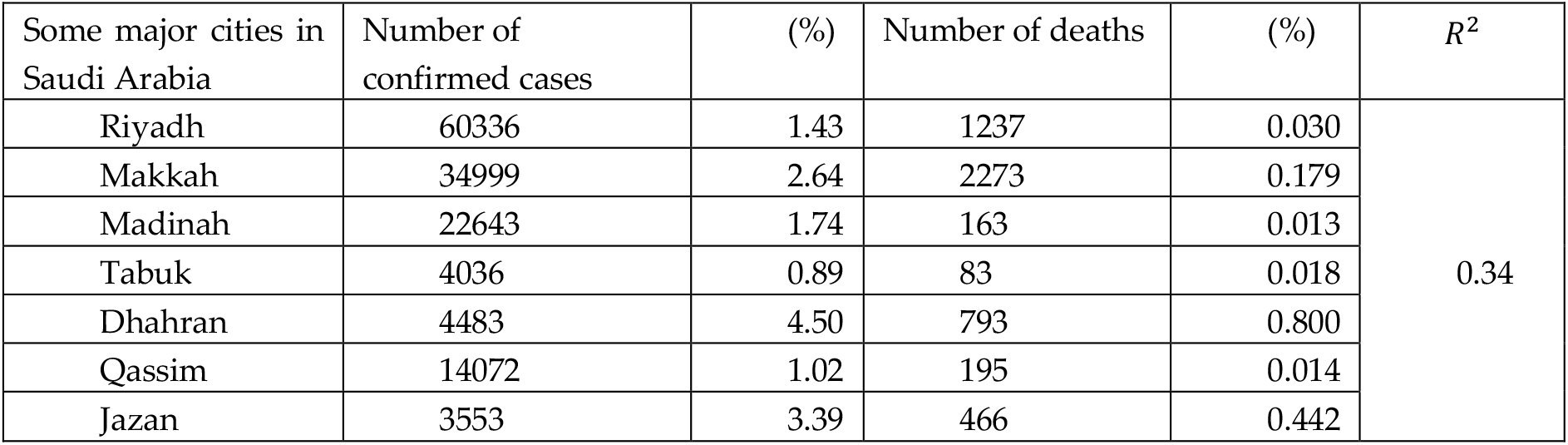
The regression analysis for the confirmed and death cases due to COVID-19 in major cities.

| Some major cities in Saudi Arabia | Number of confirmed cases | (%) | Number of deaths | (%) | $R^2$ |
| --- | --- | --- | --- | --- | --- |
| Riyadh | 60336 | 1.43 | 1237 | 0.030 | 0.34 |
| Makkah | 34999 | 2.64 | 2273 | 0.179 |  |
| Madinah | 22643 | 1.74 | 163 | 0.013 |  |
| Tabuk | 4036 | 0.89 | 83 | 0.018 |  |
| Dhahran | 4483 | 4.50 | 793 | 0.800 |  |
| Qassim | 14072 | 1.02 | 195 | 0.014 |  |
| Jazan | 3553 | 3.39 | 466 | 0.442 |  |

We collected the data related to daily confirmed, recovered and death cases due to COVID-19 released by Saudi Ministry of Health from 1^st^ May 2020 to 8^th^ January 2021. The data are arranged in a matrix form with rows representing the dates and columns represents the number of new confirmed cases, the number of cumulative confirmed cases, and the number of death cases. Table 2 shows the number cases starting from the higher infections recorded on 16^th^ June 2020. The monthly cumulative confirmed and death cases is show in Table 3. The latest updated cases in shown in Table 4.

**TABLE 2:** The number of COVID-19 cases in Saudi Arabia from 16^st^ June 2020 to 30^th^ June 2020 (*highest number of infection case).

| Dates | New Case | Cumulative confirmed cases | Recovered cases | Daily Death cases |
| --- | --- | --- | --- | --- |
| 16 June 2020 | **4919\*** | 117844 | 2122 | 39 |
| 17 June 2020 | 4757 | 122601 | 2253 | 48 |
| 18 June 2020 | 4301 | 126902 | 1849 | 45 |
| 19 June 2020 | 3941 | 130843 | 3153 | 46 |
| 20 June 2020 | 3379 | 134222 | 2213 | 37 |
| 21 June 2020 | 3393 | 137615 | 4045 | 40 |
| 22 June 2020 | 3139 | 140754 | 4710 | 39 |
| 23 June 2020 | 3123 | 143877 | 2912 | 41 |
| 24 June 2020 | 3372 | 147249 | 5085 | 41 |
| 25 June 2020 | 3938 | 108089 | 2589 | 46 |
| 26 June 2020 | 3927 | 112016 | 1657 | 37 |
| 27 June 2020 | 3989 | 116005 | 2627 | 40 |
| 28 June 2020 | 3943 | 119948 | 2363 | 48 |
| 29 June 2020 | 4387 | 124335 | 3648 | 50 |
| 30 June 2020 | 3402 | 127737 | 1994 | 49 |

**TABLE 3:**
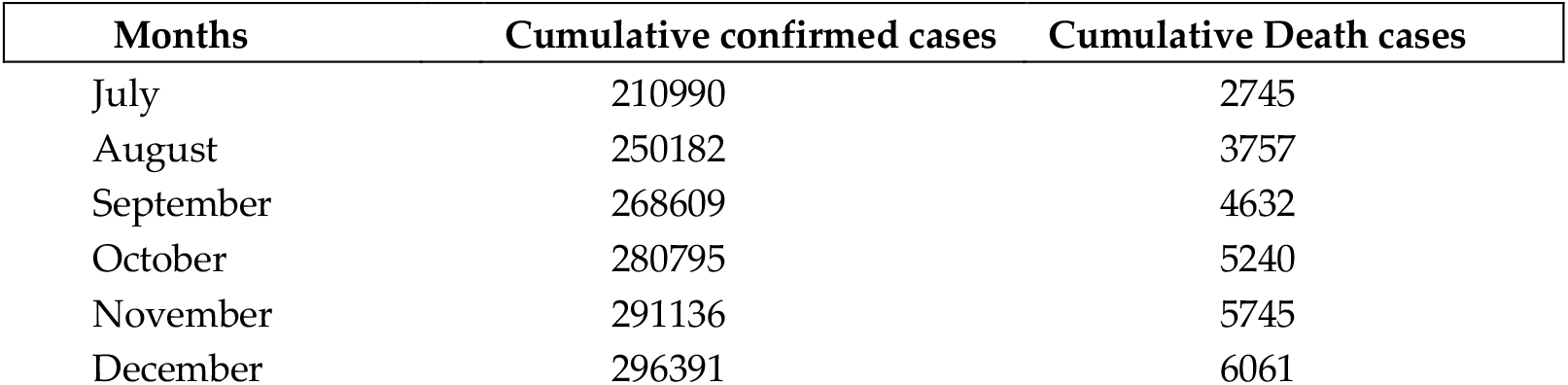
Monthly number of COVID-19 cases in Saudi Arabia from July until December 2020.

| Months | Cumulative confirmed cases | Cumulative Death cases |
| --- | --- | --- |
| July | 210990 | 2745 |
| August | 250182 | 3757 |
| September | 268609 | 4632 |
| October | 280795 | 5240 |
| November | 291136 | 5745 |
| December | 296391 | 6061 |

**TABLE 4:**
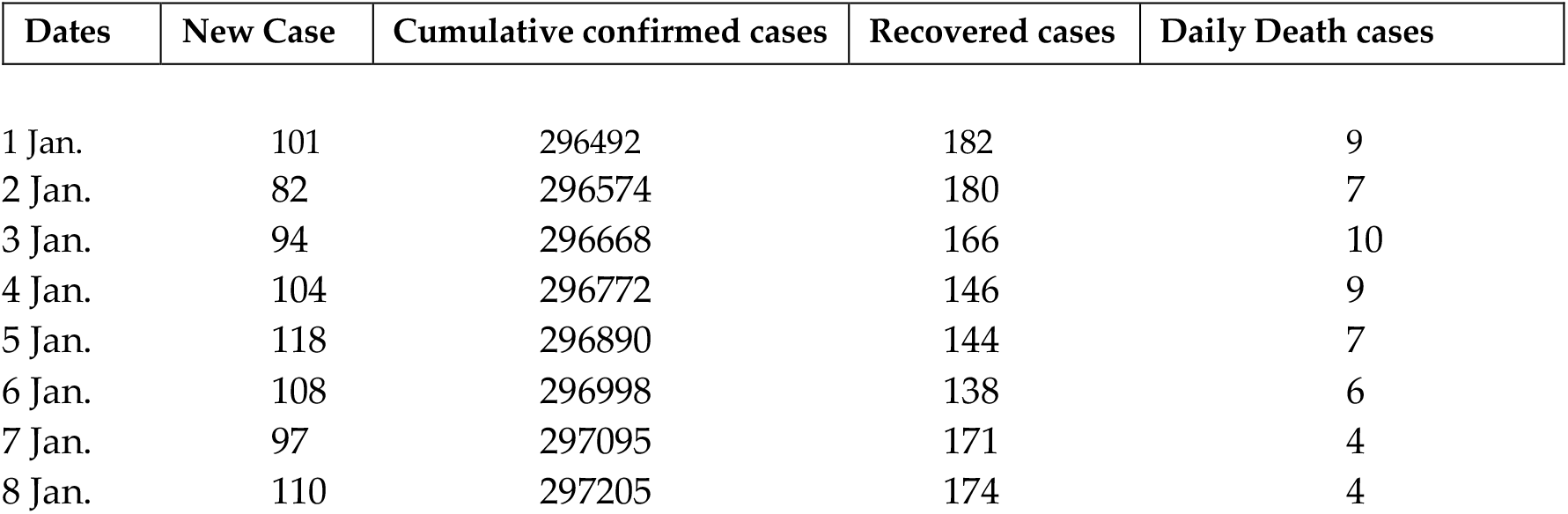
Number of COVID-19 cases in Saudi Arabia from 1^st^ January 2021 to 8^th^ January 2021.

| Dates | New Case | Cumulative confirmed cases | Recovered cases | Daily Death cases |
| --- | --- | --- | --- | --- |
| 1 Jan. | 101 | 296492 | 182 | 9 |
| 2 Jan. | 82 | 296574 | 180 | 7 |
| 3 Jan. | 94 | 296668 | 166 | 10 |
| 4 Jan. | 104 | 296772 | 146 | 9 |
| 5 Jan. | 118 | 296890 | 144 | 7 |
| 6 Jan. | 108 | 296998 | 138 | 6 |
| 7 Jan. | 97 | 297095 | 171 | 4 |
| 8 Jan. | 110 | 297205 | 174 | 4 |

From the reported cases, the number of newly confirmed case has been decreasing with higher recovery rate. This is due to effective control and prevention measures imposed by Saudi Arabia government. The following figure shows statistical analysis to interpret the real data. The analysis is performed by considering the confirmed, recovered and death cases. The Figure 2 (a) and (b), show the analysis with the daily confirmed cases as independent variable and the death cases as dependent variable. We have found that the Multiple *R*= 0.642, *R*^2^= 0.412, Adjusted *R*^2^= 0.410, Standard error = 9.452, Statistically significant *F* < 0.001 and *p* < 0.001.

**FIGURE 2:**
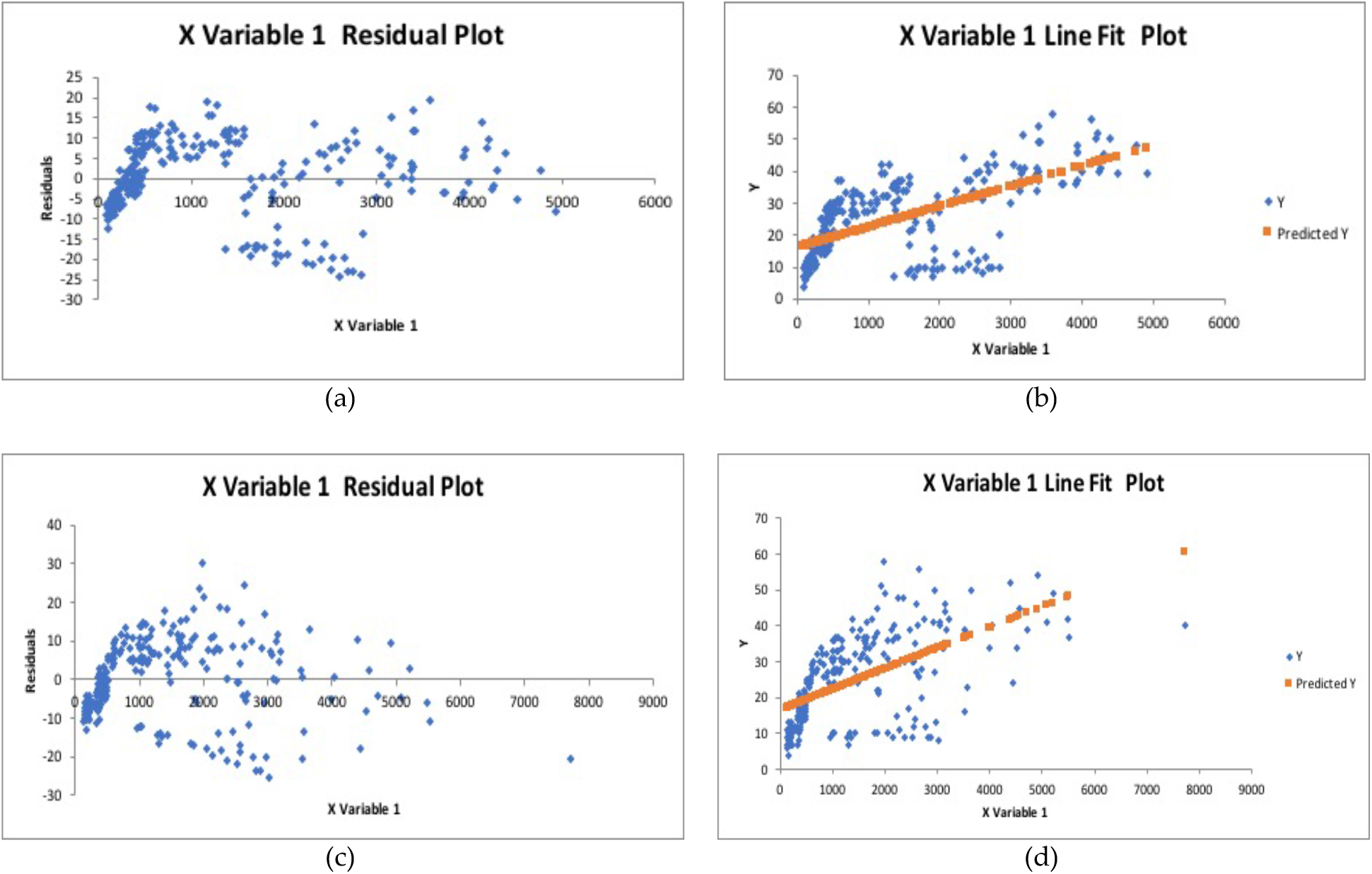
(a), (b) Statistical analysis showing residual plot and line fit plot when X = confirmed cases, Y = death cases; (c), (d) residual plot and line fit plot when X = confirmed cases, Y = recovered cases.

The Figure 2 (c) and (d), show the analysis with the daily confirmed cases as independent variable and the recovered cases as dependent variable. We have found that the multiple *R*= 0.592, *R*^2^= 0.351, Adjusted *R*^2^= 0.349, standard error = 9.93, statistically significant *F* < 0.001 and *p* < 0.001.

### 3.3 The models

We introduce long-term forecasting of the COVID-19 infection in real time by using susceptible-exposed-infected-recovered (SEIR) and Logistic growth models. We consider both the continuous and discrete cases of the logistic model equation. These models help to facilitate the prevention measure provided by the Ministry of health, Saudi Arabia by predict the number of infected people per day.

#### 3.3.1 Susceptible-exposed-infected-recovered model

The simulation of the well-known SEIR model is performed to represent the dynamic behaviors of spreading process of COID-19 epidemic in Saudi Arabia. SEIR model framework is an epidemic spreading model inspired by [22]. In epidemiology, SEIR model is one of the compartmental epidemic models widely used for characterizing the outbreak of COVID-19. In this model the spread of infection depends on the number of susceptible population and the number of infected populations. The incubation period is considered which individuals have been infected but without showing symptoms. Since the coronavirus disease has a long incubation period, it is reasonable to model the epidemic with another compartment which is Exposed humans which they are infected but not virus spreaders.

We perform simulation process to estimate the parameters of model to get the best fit on reported data of COVID-19 outbreak in Saudi Arabia. In SEIR model, we assume that the time is long-term, no vital dynamics and the population *N* size is constant. In this model, individuals are classified into four groups or compartments according to their infectious status. We classify the infection types with and without symptoms. The compartments of the model are as follows:

∘ Susceptible (*S*_*h*_): number of humans that are not infected by the virus but may catch the disease.
∘ Exposed (*E*_*h*_): number of humans that are already infected but can’t spread the virus.
∘ Infected (*I*_*h*_): number of infected humans with symptoms.
∘ Infected (*I*_*n*_): number of infected humans without symptoms.
∘ Recovered (*R*_*h*_): number of humans that are recovered from infection.

The SEIR model is

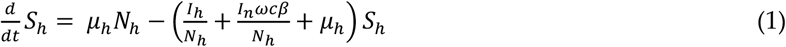

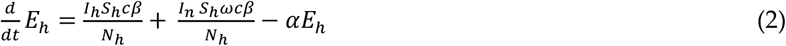

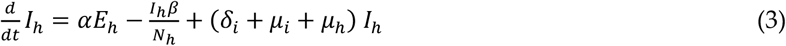

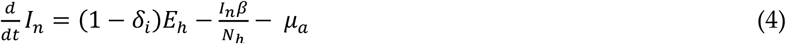

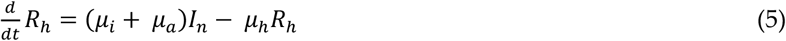

where *β*= probability of spread of the virus per contact,

*c* = rate of contact,

*ω* = transition rate of infection from people without symptoms,

*α* = transition rate of the virus of exposed individuals to the infected environment,

*δ*_*i*_ = transition rate of symptomatic infected individuals,

*μ*_*i*_ = recover rate of symptomatic infected individuals,

*μ*_*a*_ = recover rate of asymptomatic infected individuals,

*μ*_*h*_ = disease induced death rate.

The compartmental model diagram representing the SEIR is based on the dynamic behaviors of the disease, epidemiological status of the individuals and intervention measures (shown in Figure 3). We estimate the parameters of the model by using the data obtained from the confirmed cases of COVID-19 in Saudi Arabia.

**FIGURE 3:**
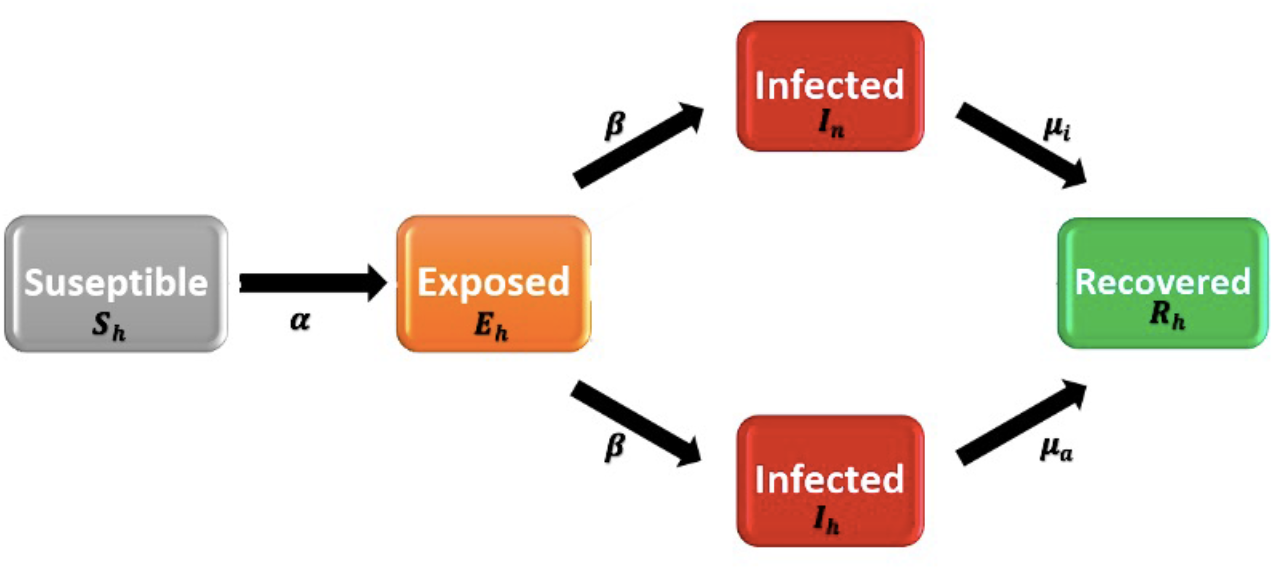
Diagram of the SEIR model adopted for simulating the novel coronavirus infection. The infection type including symptomatic and asymptomatic types.

In this model, one important parameter to understand the basic characteristics of the epidemic is the basic reproduction number (*R*_0_). It represents the average number of secondary persons in a complete susceptible population infected by a single infected person with or without symptoms during its spreading life cycle. It indicates the transmission potential of the infectious disease. The value of *R*_0_ is not certain and fixed until the outbreak is over and affected by several factors:

∘ The period of infectiousness;
∘ Probability of infection being transmitted during contact between an infected person (with or without symptoms) and susceptible person;
∘ The rate of contact between infected and susceptible person.

As the total population size is much large than the infection size. The basic reproduction number threshold is studied as an indication of transmissibility of the virus, representing the average number. We have the *R*_0_ given as follows;

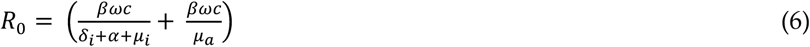

The infection of the virus is considered to be still spreading in the population when *R*_0_ > 1. And when *R*_0_ < 1, means the infection of the virus becomes decrease due to run out of susceptible and exposed individuals.

#### 2.3.2 Logistic Growth models

The logistic growth model has been widely used for predicting the population growth rate with limited resources and space. The model was originally developed by Haberman [25] and have been used to predict 2015 Ebola epidemic [27, 28]. In this model, the dynamic of epidemic is expressed as a cumulative number of cases in predicting the infection of COVID-19 in Saudi Arabia; provided that all the control measures have been implemented.

The classical logistic equation in continuous case, the epidemic can be defined as,

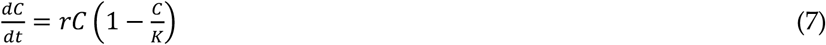

C = cumulative infection cases at time t,

r = increasing infection rate,

K = maximum infection size.

To investigate the relationship between the number of cases per day and the total number of maximum infection size, we have the following solution from (7)

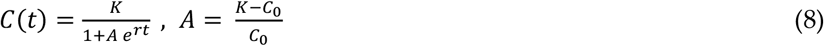

At the initial stage of the epidemic, the solution is represented by an exponential function

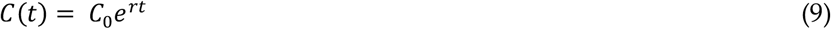

and, if graph of this equation approximates the increase in the number of cases at the initial stage, we will be able to determine the growth rate *r*. We can ignore this logistic model equation if the data do not fit with the exponential dependence.

To predict the maximum possible number of infected people is the most important characteristics. This can be predicted only at the stage of the noticeable difference the data and the exponential curve, provided the number of infected people is not small. Since the health centers, operates with the case per day, it is necessary to consider the difference logistics equation. Difference equation, which is discrete case of the logistic equation governing the epidemic as defined as follows,

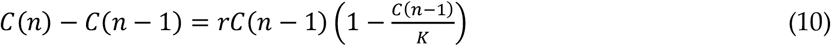

*n* = discrete time interval in days,

*C*_0_ = initial number of infections.

From equation (10), we can obtain a simple equation representing the relationship between the number of cases per day and the total number cases (maximum infection size).

## 4. Results

### 4.1 Simulation results of SEIR model

To perform the simulation model parameters, we used the empirical data of COVID-19 in Saudi Arabia which is obtained from the official website of Ministry of Health. The empirical data of confirm, recovery and death cases are collected from 1^st^ May 2020 until 8^th^ January 2021. The predicted cases from the model are compared with the actual data. We use MATLAB and MAPLE to simulate the SEIR model. The epidemic simulation graphs are as follows;

The above Figures 4, 5, and 6 show simulation of the spread of COVID-19 in Saudi Arabia, by using our modified SEIR epidemic model, considering the two types of infections symptomatic and asymptomatic. According to the figures, we can see that the infection due to the asymptotic transmission has slightly greater than the symptomatic. However, our model predicted that the rate of infection gradually decreases with time (months). In following simulation parameters (shown in Table 5) obtained from the real time data (empirical data) from Saudi Arabia are used in our model.

**TABLE 5:** Simulated parameter values.

| Parameters | $\beta$ | $c$ | $\omega$ | $\alpha$ | $\delta_i$ | $\mu_i$ | $\mu_a$ | $\mu_h$ |
| --- | --- | --- | --- | --- | --- | --- | --- | --- |
| Value | 0.095 | 14.5 | 0.009 | 0.1 | 0.13 | 0.96 | 0.94 | $1.7826 * 10^{-5}$ |

**FIGURE 4:**
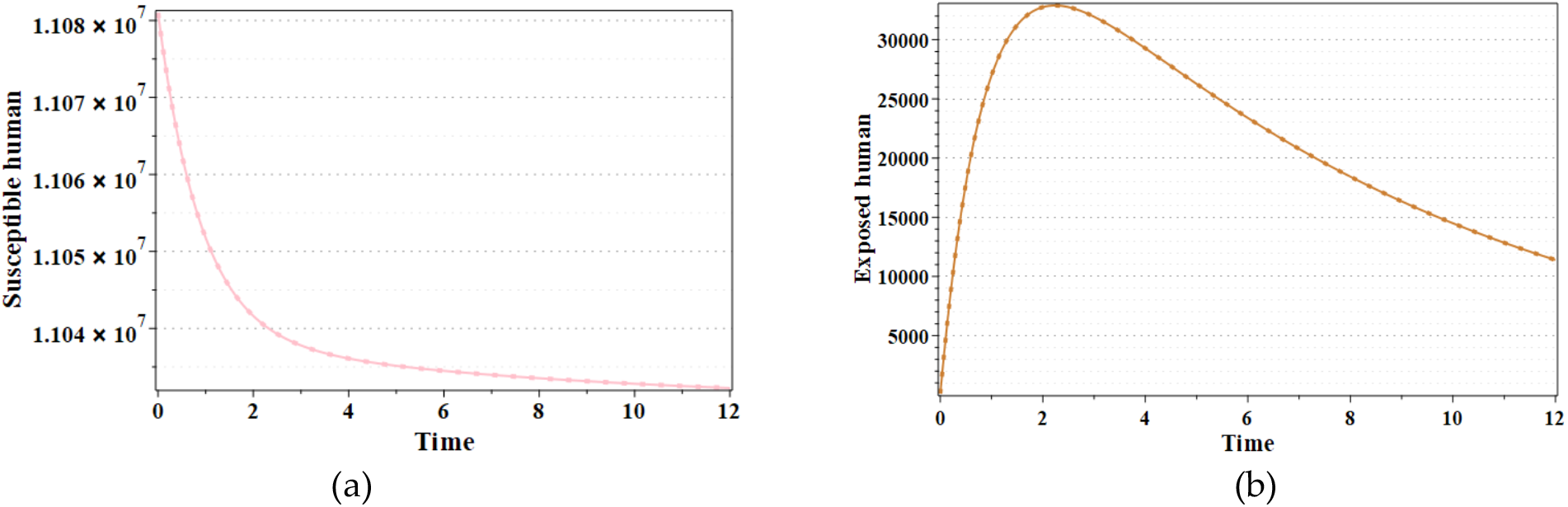
Diagram showing the (a) susceptible and (b) exposed human in Saudi Arabia prediction based on SEIR model.

**FIGURE 5:**
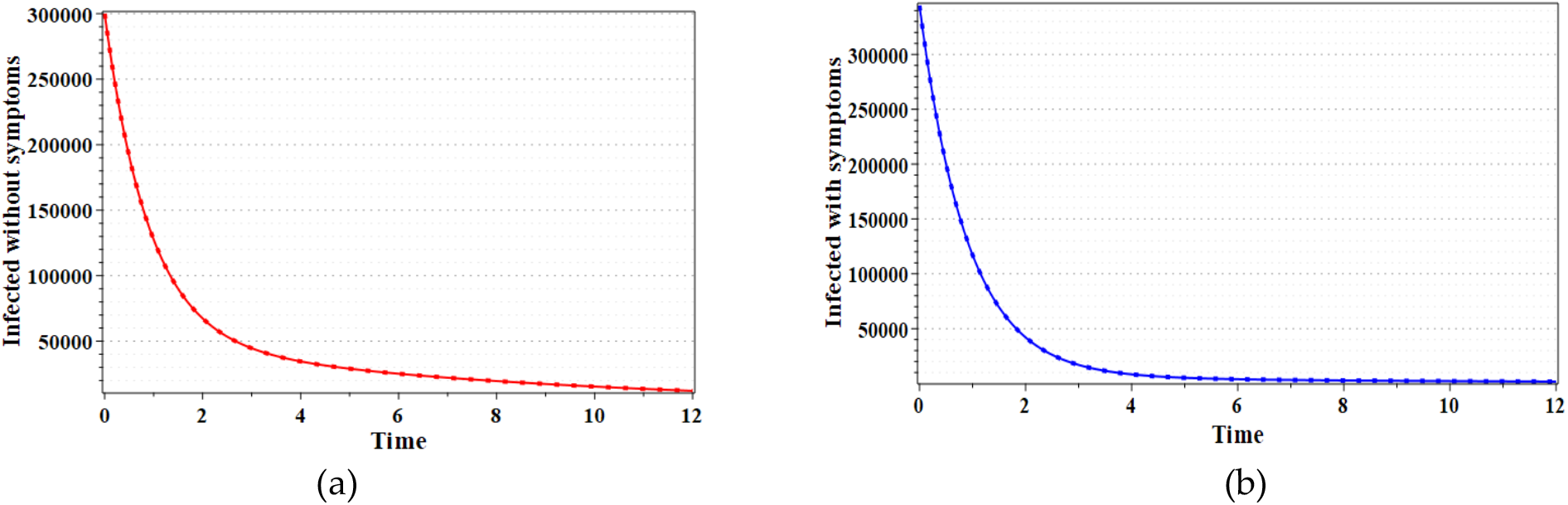
Diagram showing the (a) asymptomatic and (b) symptomatic infections predicted by SEIR model.

**FIGURE 6:**
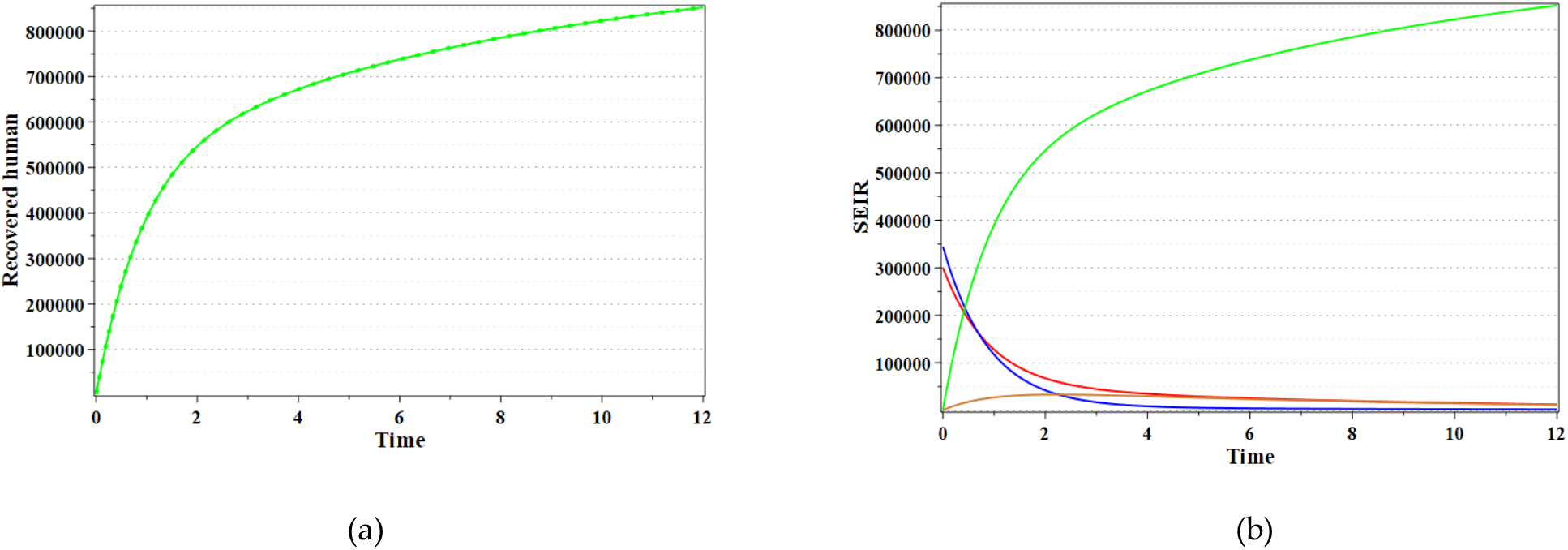
Diagram showing the (a) predicted recovered individuals in Saudi Arabia and (b) overall model of SEIR.

We used MATLAB to simulate our model equations using the above parameters and predicting the future outbreak of coronavirus in Saudi Arabia. The variation with the parameters and corresponding model behaviors is show below.

The above Figures 7 and 8 show the dynamics of the spread of coronavirus infection to human with the parameters *μ*_*i*_ and *μ*_*a*_ estimated from the actual number of cases in Saudi Arabia. From the simulated result our model predicts that the infection of the coronavirus to the environment will decrease gradually with time. According to SEIR model, the epidemic will come to its final phase with minimum infection rate at the end of February 2021.

**FIGURE 7.**
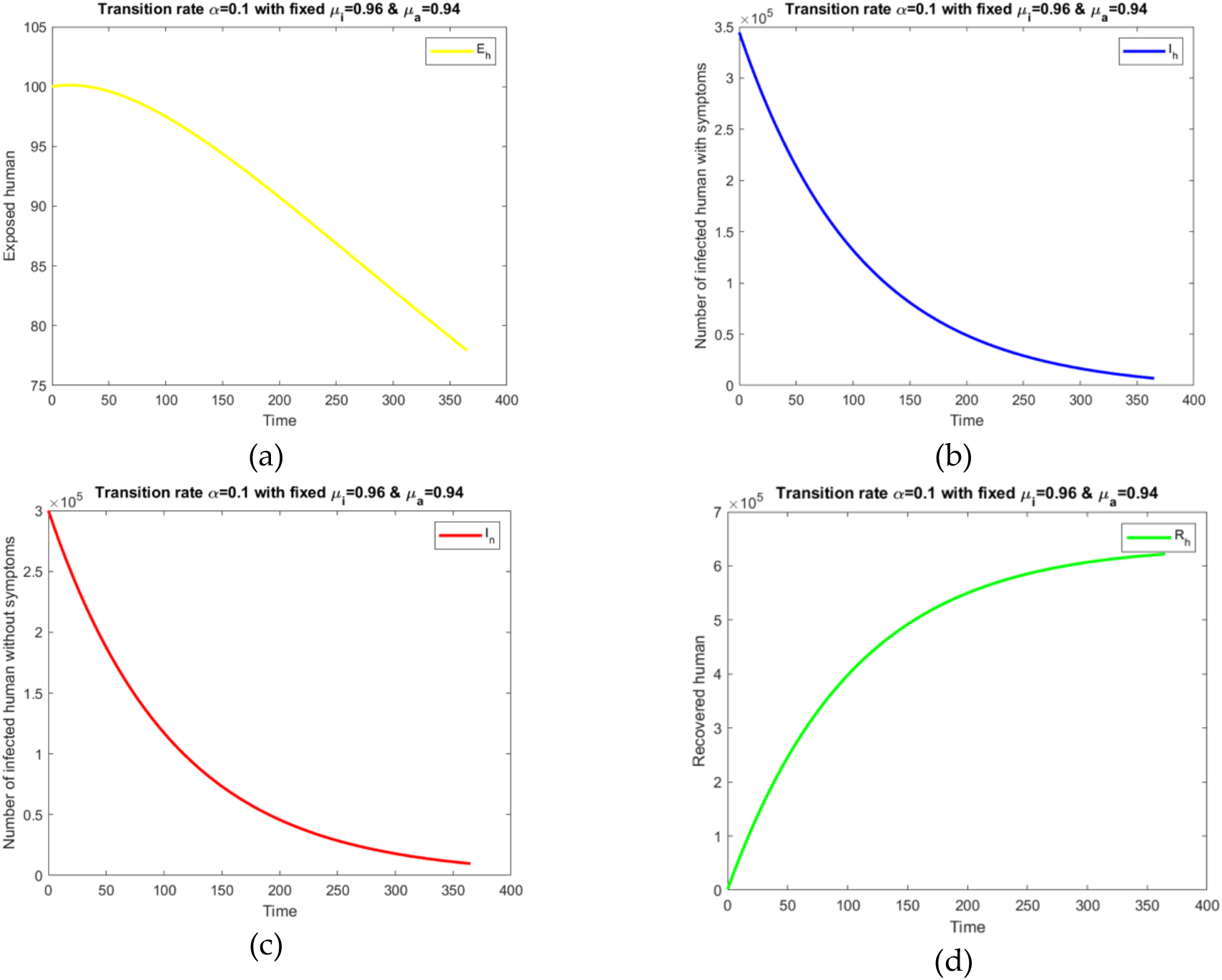
The outbreak dynamics with infection rate of the virus to the exposed individuals at the infected environment *α* = 0.1 : (a) Exposed; (b) Infected human with symptoms; (c) Infected human without symptoms; (d) Recovered human.

**FIGURE 8:**
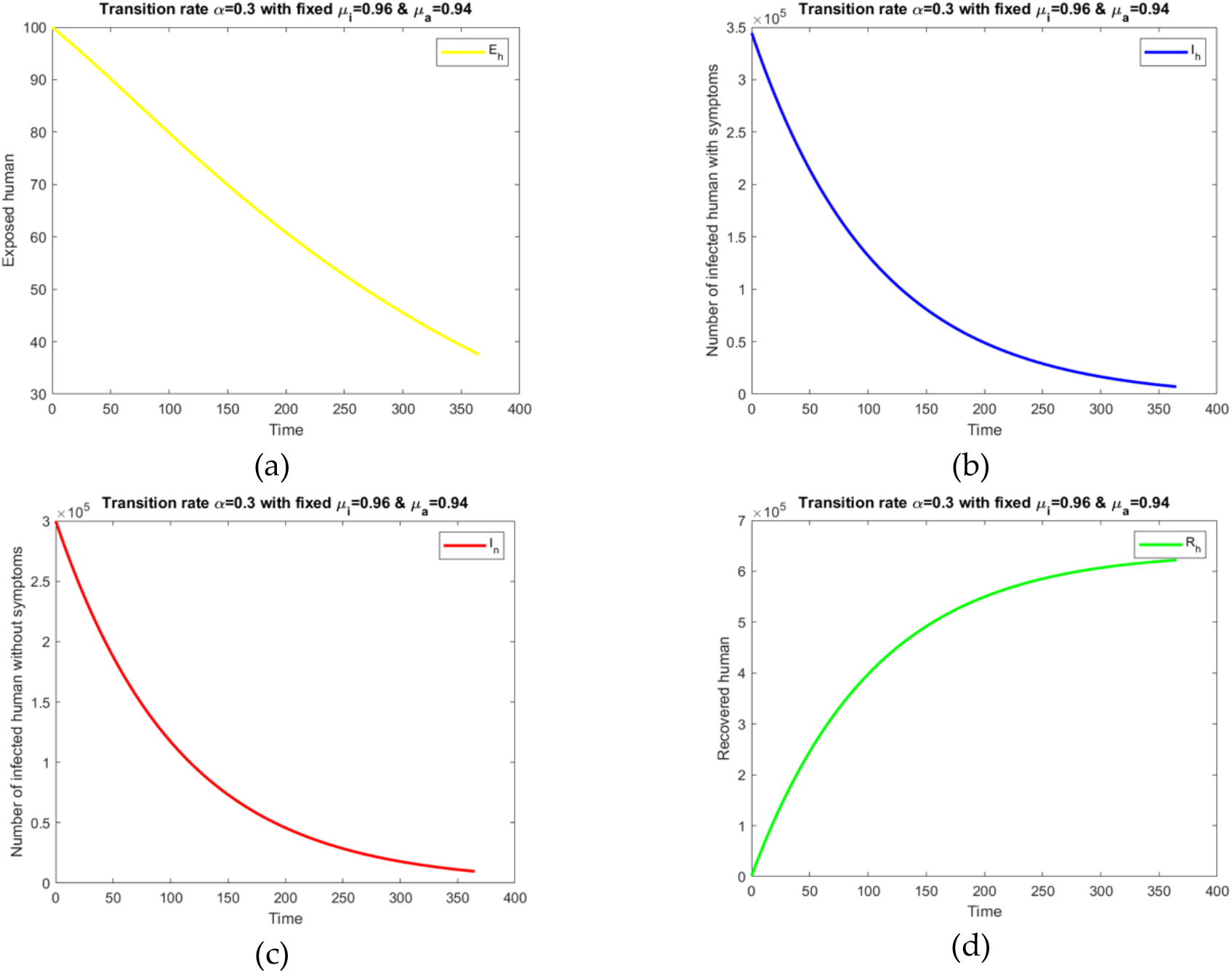
The outbreak dynamics with infection rate of the virus to the exposed individuals at the infected environment *α* = 0.3, (a) Exposed; (b) Infected human with symptoms; (c) Infected human without symptoms; (d) Recovered human.

Using the above model equations, we consider the following direction field in the various region of phase space.

From the above direction field diagrams, we can see that in Figure 9 (a), both the number of infected humans with symptoms and without symptoms heading towards a stable equilibrium point depending on the initial conditions. This states that both the infected human with and without symptoms will reduce in size the other grows towards the equilibrium point. The Figure 9 (b) shows the phase portal diagram for number of infected humans with symptoms and susceptible humans, we get negative eigenvalues and so, the equilibrium point at the *S*_*h*_ axis is stable. Hence, the susceptible humans obtain a stable stage while the infected human is minimum (almost zero infection). And the Figure 9 (c) shows the three-dimensional phase portal for susceptible, infection and recovery; the recovery humans increase with no bounds for the change in parameters related to infection and susceptible.

**FIGURE 9:**
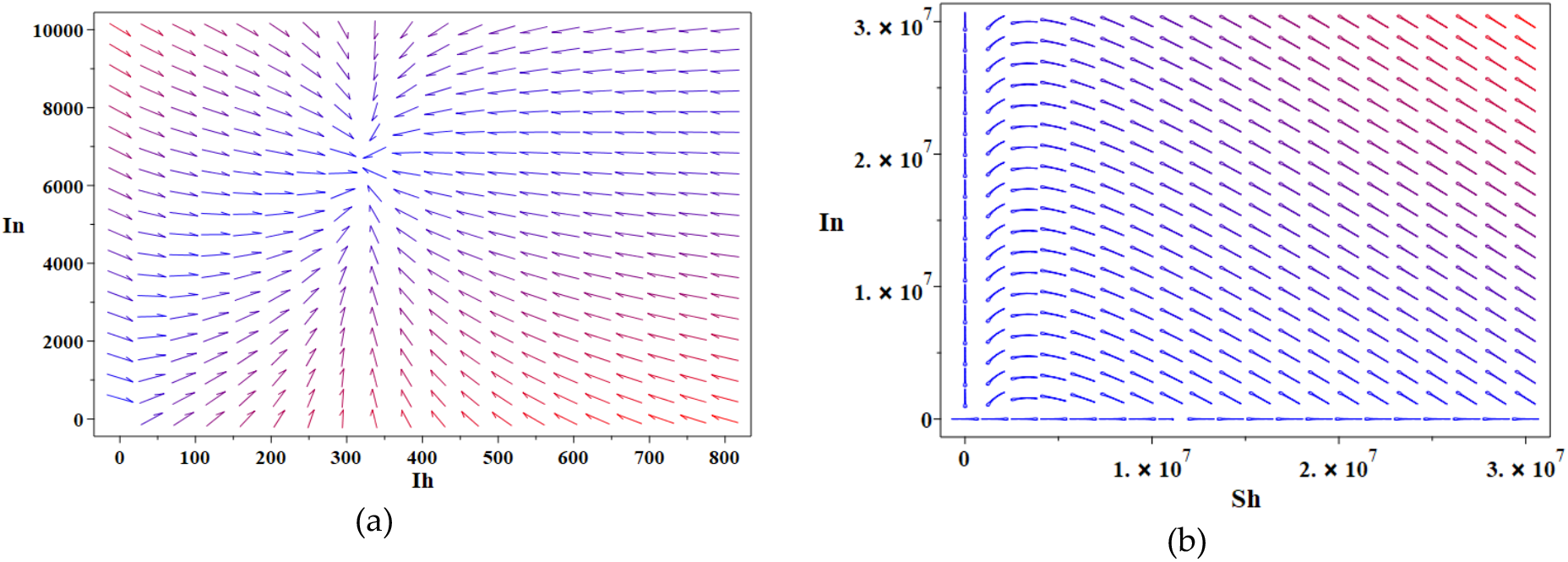
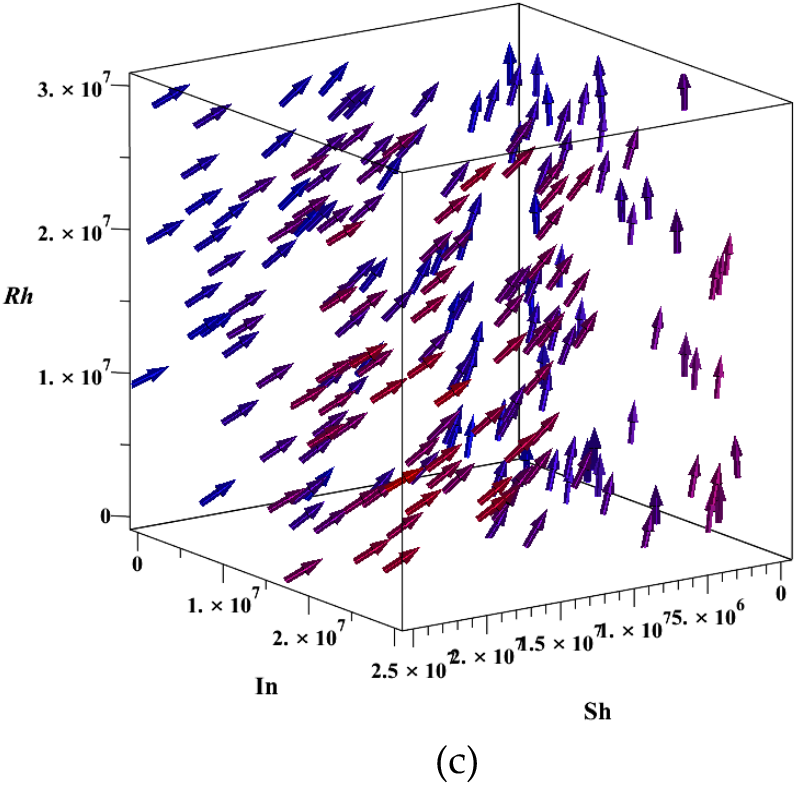
The phase portal for the SEIR model: (a) *I*_*n*_ and *I*_*h*_ plane; (b) *I*_*n*_ and *S*_*h*_ plane; (c) *R*_*h*_ and *S*_*h*_ plane at the equilibrium points.

### 4.2 Simulation results of Logistic model

We use MATLAB to simulated both continuous and discrete case of the logistic model equation. From the simulation results we compare the prediction of the cumulative infection cases with the real data. The following figures, shows the comparison of the actual and prediction of cumulative infection cases in Saudi Arabia bases on logistic growth equation.

The actual data of cumulative confirm cases from 1^st^ May 2020 are used to compare the cumulative infection case with the logistic growth model simulation and predict the future scenarios of the epidemic. We use exact and discrete case of the logistics growth models to predict the cumulative infection cases in Saudi Arabia. In Figure 10 (a), (b) and (c) shows predicted cumulative infection cases with infection rate 0.015 for 360, 400 and 460 days respectively. The simulation is repeated with the infection rate 0.016 for 360, 400 and 460 days shown in Figure 10 (b), (d) and (f) respectively. These models both discrete and continuous cases tend to underestimate the maximum infection size K, and thus could serve as lower bounds of future prediction. The logistic growth model provides accurate results in estimating the final size of the epidemic in Saudi Arabia.

**FIGURE 10:**
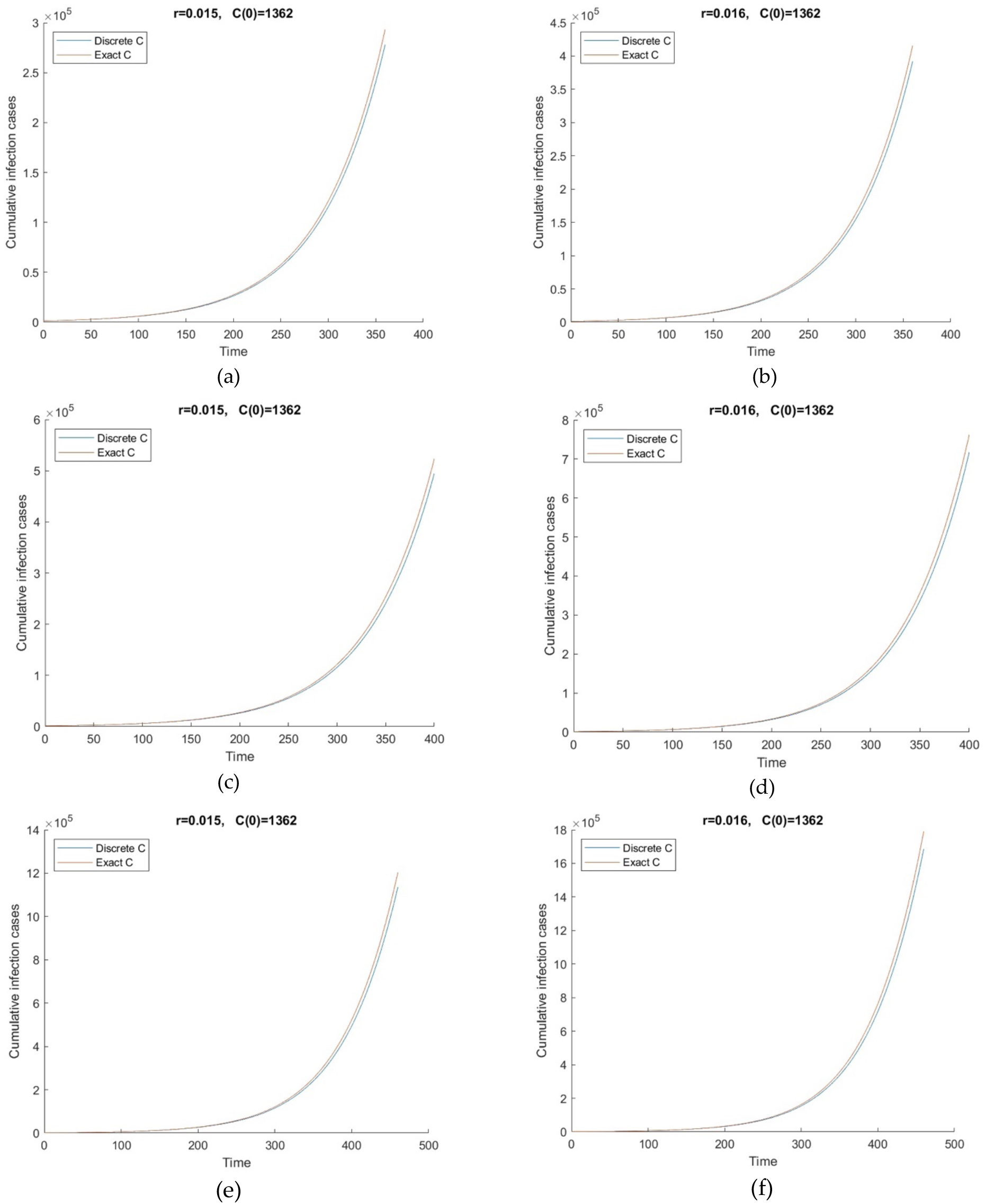
The prediction of cumulative infection cases in Saudi Arabia based on logistic growth rate models with infection rate is 0.015 for (a) 360 days, (c) 400 days, (e) 460 days; and with infection rate is 0.016 for (b) 360 days, (d) 400 days, (f) 460 days.

## 5. Discussion

Our proposed SEIR model considering both the infection types with and without symptoms is suitable for slow transmission of COVID-19 in Saudi Arabia. The parameters are simulated from the real data and found different with different scenarios. Moreover, the logistic growth model is used to predict future dynamics of the outbreak in Saudi Arabia.

COVID-19 infection is an important issue concerning the global health and has infected wide area of the world’s population. In this study, we investigated the biological and epidemiological trends in the prevalence and mortality due to outbreak of novel coronavirus infection. The infection of COVID-19 is expanding to over 220 countries and territories. It has infected 89,858,494 people, and has caused 1,930,829 (2.14%) death during the period December 2019 to 8^th^ January 2021.

The infection of COVID-19 is rapidly transmitted during the months June, July and August. The spreading phase of coronavirus has gradually slow down due to various control measure such as lockdown and limitation of traveling. It is highly contagious due to its biological characteristics [31]. The infected people can transmit the disease before they show clinical symptoms and it is contagious. This is one of the main reasons that the virus could swiftly spread from Wuhan city, China to rest of the world and in Saudi Arabia. However, the Ministry of Health (MOH), Saudi Arabia has taken various precautionary measures to control the spread of COVID-19. Some of the preventive measures taken by MOH are as follows; (1) providing COVID center help line number 937 and suspension of entry into Saudi Arabia for Umrah, (2) ceasing to issue visas individuals coming from countries that had already been affected by COVID-19, (3) banning citizen of Gulf countries entering two holy cities Makkah and Almadinah, (4) more than twenty five hospitals all over Saudi Arabia were addressed COVID-19 cases, (5) massive campaigns for awareness in public places and educational organizations [32]. All these preventive and precautions played an important role in limiting the spread of COVID-19 infection in Saudi Arabia. Also, the number of death rate as compare with the other countries is very low, which might be due to good care of infected patients in hospitals in Saudi Arabia.

Recently, the outbreak of the new mutated coronavirus variant has been identified in UK. The health secretary of UK has mentioned that new variant of COVID-19 may be driving infection in south east region. One of the most significant mutation of this virus is in the spike protein that may result in becoming more infectious and spreading rapidly between people [29].

The three main characteristics that comes together making the new coronavirus variant SARS-CoV-2 easier to spread are as follows;

∘ rapidly replacing another version of the virus,
∘ mutations that affect part of the virus,
∘ some of those mutations increase the ability of the virus to infect cells [30].

Over time, as more such mutations occur, the existing vaccine may need to adjusted accordingly. However, SARS-CoV-2 doesn’t not mutate quickly as flu virus, and the vaccines that have so far proved effective in trails is an improvement. There is no detection of new strain of the coronavirus in Saudi Arabia. The Saudi health representative also stated that the mutated coronavirus variant did not change the way the virus affects human [6] (accessed on 27 December 2020).

Considering the present scenario, adopting the precaution measures and travel limitations with the other countries, we have predicted the possible number of cumulative infection cases in Saudi Arabia. The Table 6, shows estimated prediction for future epidemic trends using logistic growth rate model. The graphical representation of predicting the epidemic is shown in Figure 8. We can observe that the prediction using our models are less significant difference with the real data. The prediction number with the exact solution of the continuous logistic growth model shows more accurate and very close to the cumulative case of the real data. Using the exact solution of the logistic model, the estimation of the cumulative coronavirus cases around 360 days is 293544 which is close to the real cumulative case 296492. Further, we have estimated the possible cumulative infection cases for 400 and 460 days.

**TABLE 6:** Outbreak prediction of cumulative confirmed cases.

| Model | 360 days | 400 days | 460 days |
| --- | --- | --- | --- |
| Predicted by logistic model ($r = 0.015$) | Discrete: 278338<br>Exact: 293544 | Discrete: 494863<br>Exact: 523390 | Discrete: 1136294<br>Exact: 1203734 |
| Predicted by logistic model ($r = 0.016$) | Discrete: 392238<br>Exact: 415936 | Discrete: 717755<br>Exact: 762952 | Discrete: 1687537<br>Exact: 1792251 |
| Real data (cumulative) | 296492 |  |  |

In Table 7, prediction of the possible number of daily infection cases (Active cases) in Saudi Arabia is shown using the SEIR model. Simulation of the outbreak prediction is performed for three phases 360, 400 and 460 days. The prediction of the daily infection cases using the SEIR model considering the type of infection with symptoms and without symptoms is simulated and its graphical representation is shown in the Appendix A. From the simulation process, the SEIR model with infection types has higher prediction compare with the real data on daily active cases. However, the estimation of the infection nature has reduced over the period of time. The infection rate of the virus from the asymptomatic individuals are seem to have higher than the asymptomatic individuals. It is suggested that the asymptomatic people should be identified to contain the spread of the disease.

**TABLE 7:** Outbreak prediction of confirmed cases per day.

| Model | 360 days | 400 days | 460 days |
| --- | --- | --- | --- |
| Predicted by SEIR model ($r = 0.015$) | Infection ($I_h$): 10358<br>Infection ($I_n$): 7526 | Infection ($I_h$): 7075<br>Infection ($I_n$): 3668 | Infection ($I_h$): 4064<br>Infection ($I_n$): 111 |
| Predicted by SEIR model ($r = 0.016$) | Infection ($I_h$): 10262<br>Infection ($I_n$): 7414 | Infection ($I_h$): 7074<br>Infection ($I_n$): 3669 | Infection ($I_h$): 4063<br>Infection ($I_n$): 112 |
| Real data (Active case) | 2772 |  |  |

Our future research directions would focus on extension of the proposed models with availability of vaccination and more suitable with the mutated SARS-CoV-2.

## 5. Conclusions

The aim of this research work is to model and simulate the spread of COVID-19 in Saudi Arabia using compartmental epidemic models. We study the dynamic nature of the coronavirus outbreak in Saudi Arabia for the reported cases from 1^st^ May 2020 to 8^th^ January 2021 using mathematical modeling and simulation. We used both the SEIR and logistic growth models to simulate the further spread of COVID- 19. The SEIR model has been extended by considering the infection types both symptomatic and asymptomatic. According to the model, the asymptomatic infected individuals have higher chance of spreading the virus. Also, we adopted logistic growth model for both discrete and continuous time to understand the transmission dynamics of the novel coronavirus in Saudi Arabia. The models predict the cumulative confirmed cases and daily active cases through simulation process. However, the logistic growth model provides a very good prediction close to the real number of cases. To describe the daily active cases, the deterministic logistic model is more appropriate. Both the models have their own advantages in predicting the future number of cases with SEIR model has higher than the logistics model.

In conclusion, we would like to say that our models predicted the future nature of the outbreak and assess the effectiveness of Saudi Health Ministry control measures on infection. Furthermore, according to our model predictions, the future outbreak under the strict control measures, is expected to its minimum during the month of February 2021.

## Data Availability

In our study, the information on epidemiological trends and current situation of coronavirus COVID 19 infection is taken from the World Health Organization, Worldmeter-Coronavirus and Ministry of Health, Covid19 Command and Control Center CCC (free accessible data).

## Author Contributions

All authors have read and agreed to the published version of the manuscript.

## Funding

This study was funded by Scientific Research Ethics Committee at Jazan University, Saudi Arabia under the grand number (Cov19-38).

## Acknowledgments

The authors would like to thank Scientific Research Ethics Committee, Jazan University, Ministry of Higher Education, Saudi Arabia, for financial support of this research.

## Conflicts of Interest

The authors declare no conflict of interest.

## Appendix A

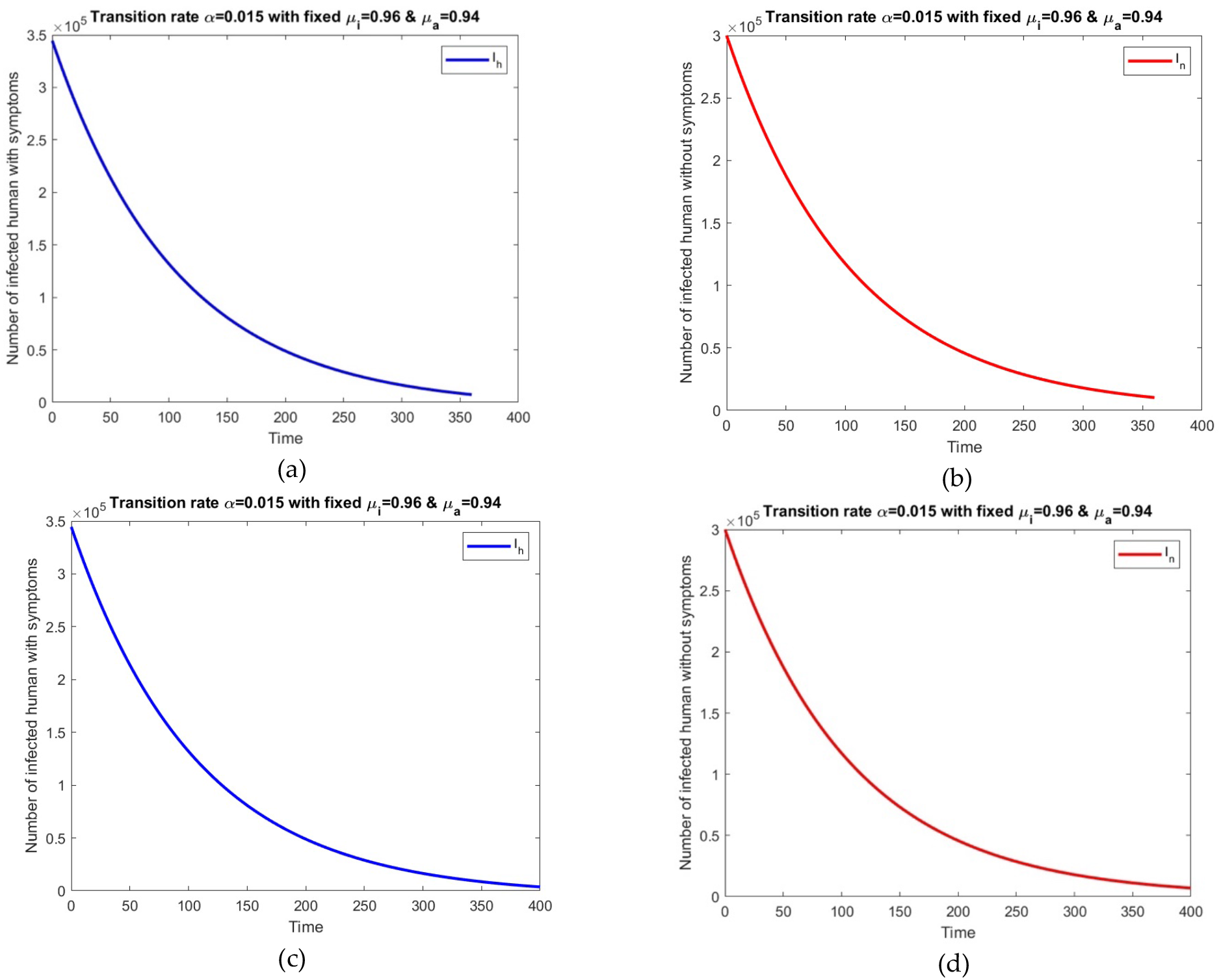
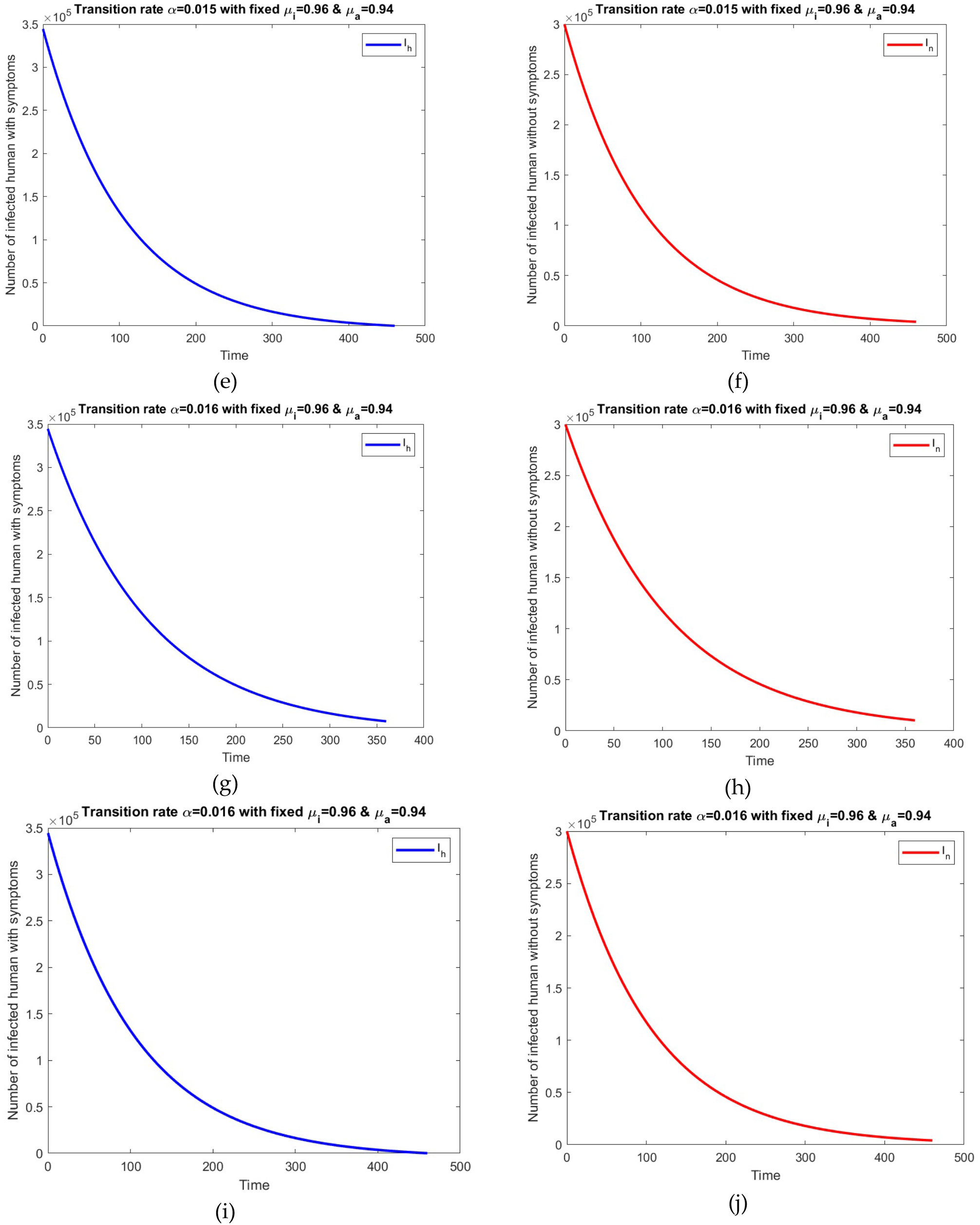

## Notes

### Competing Interest Statement

The authors have declared no competing interest.

### Funding Statement

This study was approved and funded by Scientific Research Ethics Committee at Jazan University, Saudi Arabia.

### Author Declarations

The study of this research work is conducted in the department of mathematics, college of science, Jazan University, Jazan, Saudi Arabia. Only free accessible date sources were used, so ethical approval is not required. This study is part of the research project with grant number (Cov19-38), funded by Scientific Research Ethics Committee at Jazan University, Saudi Arabia.

